# Shedding of infectious SARS-CoV-2 from airways in hospitalized COVID-19 patients in relation to serum antibody responses

**DOI:** 10.1101/2020.09.11.20191940

**Authors:** Hedvig Glans, Sara Gredmark-Russ, Mikaela Olausson, Sara Falck-Jones, Renata Varnaite, Wanda Christ, Kimia T. Maleki, Maria Lind Karlberg, Sandra Broddesson, Ryan Falck-Jones, Max Bell, Niclas Johansson, Anna Färnert, Anna Smed-Sörensen, Jonas Klingström, Andreas Bråve

**Author notes:** **Corresponding author**: Hedvig Glans.

## Abstract

To understand the risk of transmission of SARS-CoV-2 in hospitalized COVID-19 patients we simultaneously assessed the presence of SARS-CoV-2 RNA, live infectious virus in the airways, and virus-specific IgG and neutralizing antibodies in sera in 36 hospitalized COVID-19 patients. SARS-CoV-2 could be cultured from four patients, all with low or undetectable antibody response. Our data suggests that the level of SARS-CoV-2 antibodies may correlate to risk for shedding live SARS-CoV-2 virus in hospitalized COVID-19 patients.

## Introduction

It is important to better understand the risk of severe acute respiratory syndrome coronavirus 2 (SARS-CoV-2) transmission from coronavirus disease 2019 (COVID-19) patients. Many studies report PCR-detectable SARS-CoV-2-RNA in the airways over time (1–3), but little has been reported on shedding of infectious viruses from the airways in hospitalized COVID-19 patients. In the setting of mild COVID-19 infection, shedding of infectious virus has been described during the first week of symptoms (4). No viable virus could be isolated after 8 days, coinciding with the appearance of detectable antibodies to SARS-CoV-2 (5). In a larger study, with patients ranging from asymptomatic to severe, SARS-CoV-2 culture-positive airway samples declined to 6% at day 10 (6). In a cohort with 129 severely ill COVID-19 patients, the median duration of shedding of infectious virus was 8 days after symptom onset, with infectious virus found up to 20 days after symptom onset (7). Determinants for the detection of infectious SARS-CoV-2 were either viral loads above 7 log10 RNA copies/mL in airways or neutralizing antibody titres below 20 in sera (7). Even if there is one case report on a patient with SARS-CoV-2 shedding for 18 days even after seroconversion at day 10 (8), antibodies may be of importance in judging contagiousness.

The aim of our study was to evaluate if infectious SARS-CoV-2 could be cultured from nasopharyngeal and sputum samples in hospitalized patients with symptoms exceeding 8 days and to evaluate if the SARS-CoV-2-specific antibody responses in serum correlated with shedding of infectious virus.

## Results

Thirty-six COVID-19 patients were enrolled in this study, the majority were men (n=25, 69%), and the median age was 60.5 years (range 25-77 years) (Table 1). The majority of the patients had an underlying risk factor for severe COVID-19 infection (Table 1) (9). Thirty-three (92%) patients received supplemental oxygen treatment. Fourteen (39%) patients were treated in the intensive care unit, of which 9 were treated with mechanical ventilation. Fourteen patients had received immunomodulatory treatment, and four had received remdesivir, prior to inclusion in the study (Table 1).

**Table 1.**
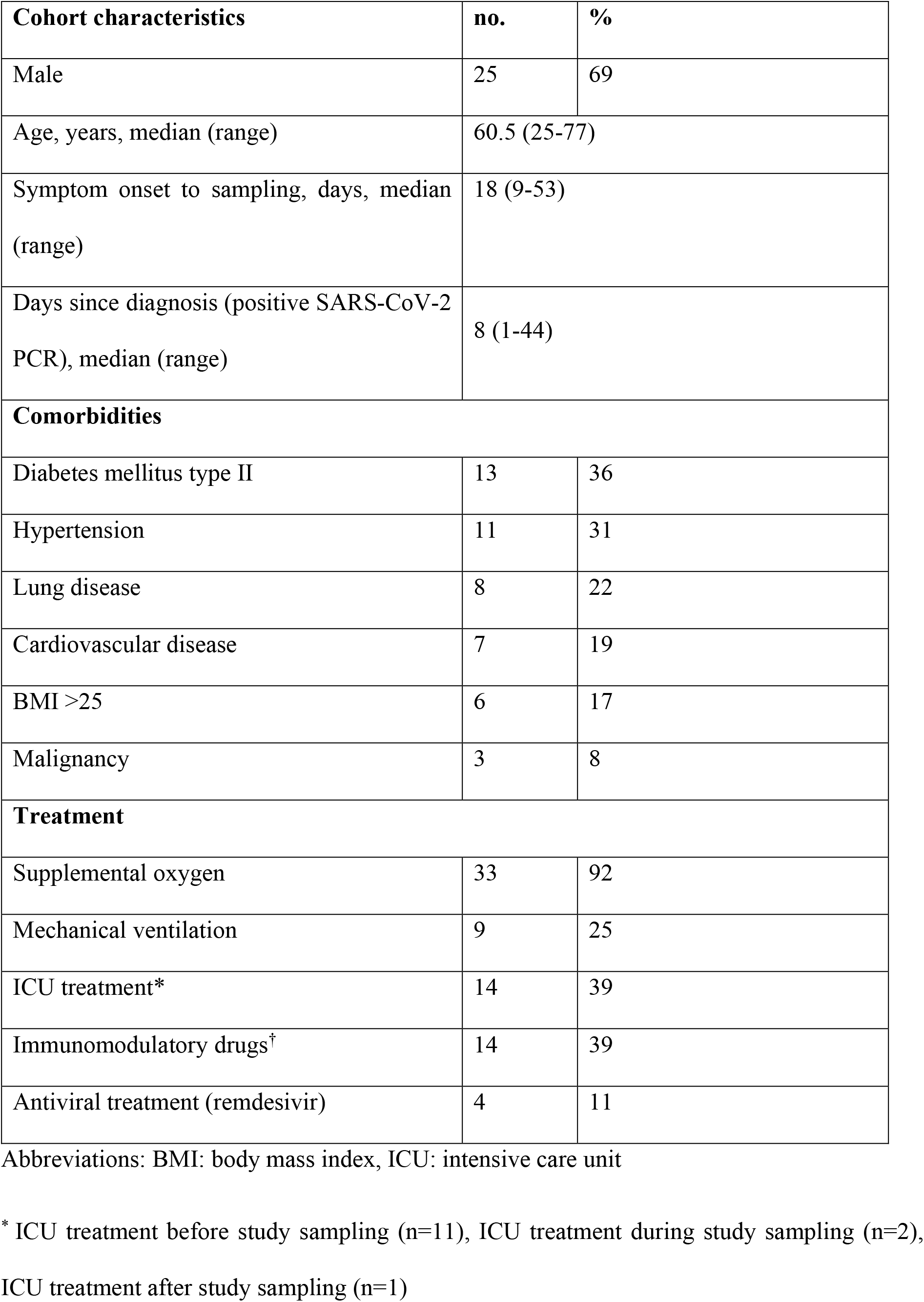
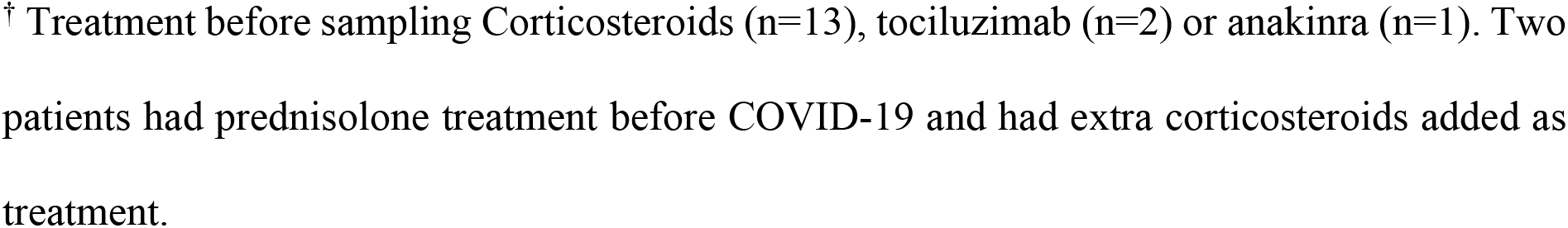
COVID-19 patient characteristics (n = 36).

| Cohort characteristics | no. | % |
| --- | --- | --- |
| Male | 25 | 69 |
| Age, years, median (range) | 60.5 (25-77) |  |
| Symptom onset to sampling, days, median (range) | 18 (9-53) |  |
| Days since diagnosis (positive SARS-CoV-2 PCR), median (range) | 8 (1-44) |  |
| Comorbidities |  |  |
| Diabetes mellitus type II | 13 | 36 |
| Hypertension | 11 | 31 |
| Lung disease | 8 | 22 |
| Cardiovascular disease | 7 | 19 |
| BMI >25 | 6 | 17 |
| Malignancy | 3 | 8 |
| Treatment |  |  |
| Supplemental oxygen | 33 | 92 |
| Mechanical ventilation | 9 | 25 |
| ICU treatment* | 14 | 39 |
| Immunomodulatory drugs† | 14 | 39 |
| Antiviral treatment (remdesivir) | 4 | 11 |

Abbreviations: BMI: body mass index, ICU: intensive care unit
\* ICU treatment before study sampling (n=11), ICU treatment during study sampling (n=2),
ICU treatment after study sampling (n=1)
<sup>†</sup> Treatment before sampling Corticosteroids (n=13), tocilizumab (n=2) or anakinra (n=1). Two patients had prednisolone treatment before COVID-19 and had extra corticosteroids added as treatment.

Samples from the patients were collected at a median of 18 days (range 9-53 days) after self-reported onset of symptoms, and at a median of 8 days (range 1-44 days) after their positive diagnostic sample (Table 1).

Twenty-three (64%) patients had SARS-CoV-2 RNA detected by PCR in airway samples, of which 14 were positive in nasopharynx and 18 in sputum (Supplementary Table 1).

SARS-CoV-2 could be cultured from airway samples in four patients (11% of all patients, 17.3% of PCR-positive patients) (Table 2). All four patients had underlying diseases potentially increasing the risk of severe COVID-19 (9).Three of the patients had been symptomatic for 9-11 days before sampling. The fourth patient, with a haematological malignancy and leukopenia during hospitalization, still had infectious SARS-CoV-2 on day 16 after onset of symptoms.

**Table 2.** Antibody and virus levels in SARS-CoV-2 culture-positive patients

| Patient ID* | SARS-CoV-2 PCR (Ct value) |  |  | SARS-CoV-2 culture |  | Anti-SARS-CoV-2 IgG titer | Neutralizing antibody titer | Days after onset of symptom |
| --- | --- | --- | --- | --- | --- | --- | --- | --- |
|  | NPH | Sputum | Serum | NPH | Sputum |  |  |  |
| 1 | 19 | 21 | neg | pos | pos | <20 | <10 | 16 |
| 2 | 24 | 28 | neg | pos | neg | <20 | <10 | 9 |
| 3<sup>†</sup> | 25 | 19 | neg | pos | neg | 40 | <10 | 9 |
| 4<sup>†</sup> | 26 | 20 | neg | pos | neg | 40 | 10 | 11 |

\*Comorbidities: patient 1 - follicular lymphoma, diabetes mellitus II, chronic obstructive pulmonary disease, ST elevation myocardial infarction (STEMI); patient 2 - diabetes mellitus II, hypertension, chronic kidney failure; patient 3 - asthma, hypothyroidism; patient 4 - asthma, obesity.
<sup>†</sup> Betametason treatment (4 mg) within 24 hours before sampling.
Abbreviations: neg: negative, NPH: nasopharynx, pos: positive

Three (8.3%) patients had detectable but low levels of SARS-CoV-2 RNA in blood (Supplementary Table 1), however, none of the patients were culture positive for SARS-CoV-2 in the respiratory samples.

All but two patients had detectable levels of SARS-CoV-2-specific immunoglobulin G (IgG) in serum, with titers ranging from 1:40 to 1:2560. Neutralizing antibodies towards SARS-CoV-2 were detected in 33 of the 36 patients (Supplementary Table 1). The positive control, sampled at day 5 after onset of symptoms, had no detectable SARS-CoV-2-specific antibodies, but was positive for infectious virus in the airways (data not shown).

There was a strong positive correlation between total SARS-CoV-2-specific IgG titers and neutralizing antibody titers (Spearman’s correlation: r_s_ = 0.689, p < 0.001) (Figure 1A).

**Figure 1.**
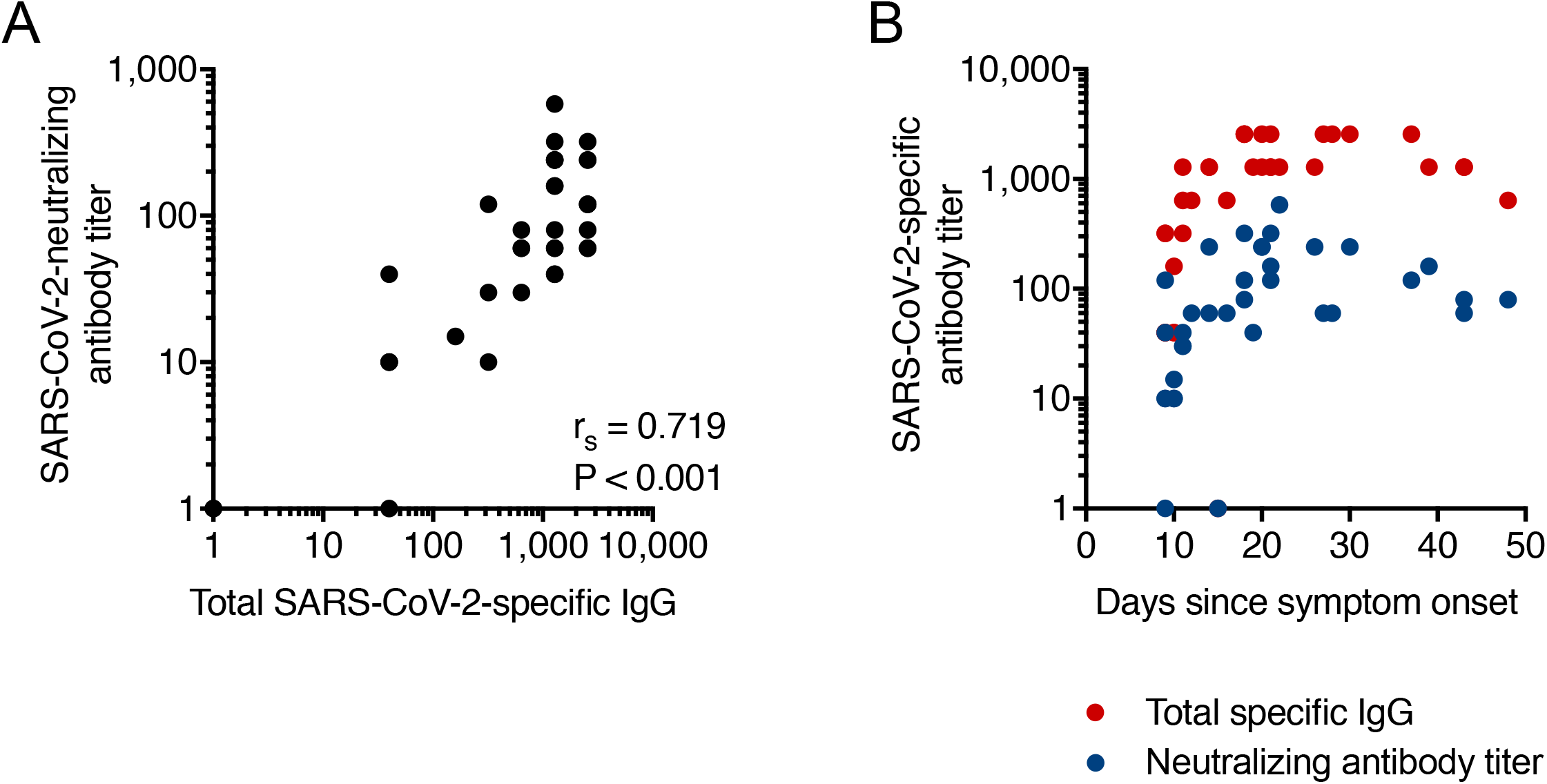
A. Correlation between anti-SARS-CoV-2 IgG titers and neutralizing antibodies titers. B. SARS-CoV-2-specific and neutralizing antibody levels in patients plotted on a timeline.

Patients sampled earlier after symptom onset, showed a variation in IgG titers and neutralizing antibodies (Figure 1B and Supplementary Table 1). Patients with IF titers equal to or below 1:40 either displayed no detectable, or low levels of neutralizing antibodies (Supplementary Table 1). Whereas, the distribution of the levels of SARS-CoV-2 IgG and neutralizing antibody titers within the patients are similar, and with a general higher level of antibodies and neutralizing titer in patients sampled during later times (Figure 1B), all four patients that shedded infectious SARS-CoV-2 had low or no detectable IgG titers and neutralizing antibody titers (Table 2 and Table 3).

**Table 3.**
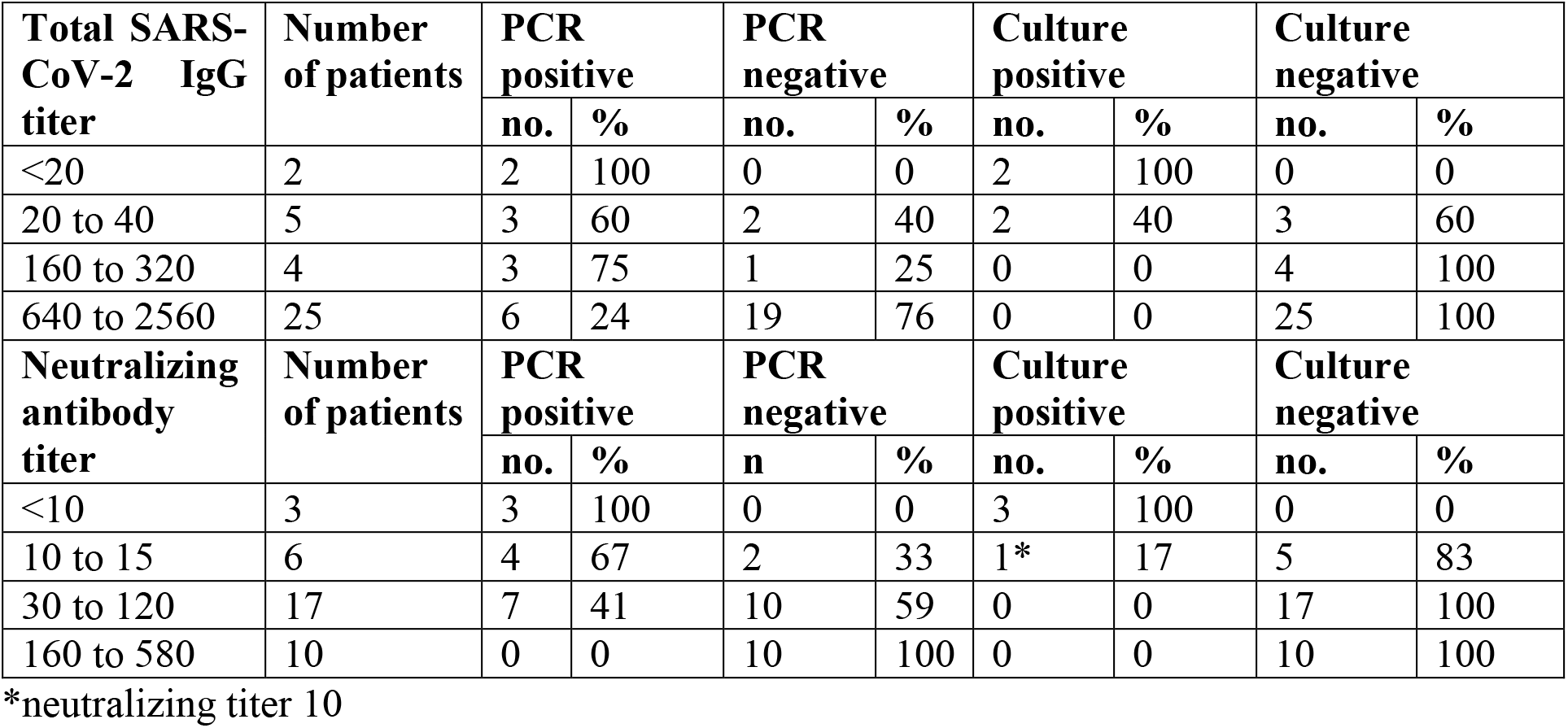
SARS-CoV-2-specific antibody titers in relation to detectable SARS-CoV-2 RNA and infectious SARS-CoV-2 in nasopharynx in all COVID-19 patients (n = 36).

## Discussion

Viable SARS-CoV-2 were isolated from four patients. We could not detect viable SARS-CoV-2 virus from patients with SARS-CoV-2-specific IgG titers above 1:40, or with neutralizing antibodies titers above 1:10. Seroconversion, with detectable neutralizing antibodies, was observed in all 32 patients from whom no viable SARS-CoV-2 virus could be isolated from the respiratory tract. In addition, no infectious virus could be isolated from the patients that were SARS-CoV-2 RNA PCR negative.

In patients with a shorter symptom duration, there was a large variation in total specific IgG and neutralizing antibody titers. Three of the culture-positive patients were sampled within 11 days after the onset of symptoms. However, the fourth patient with culturable SARS-CoV-2, an immunocompromised patient, was sampled at day 16 after the onset of symptoms, showing that infectious virus can be shed rather late during COVID-19 in immunocompromised patients.

Similar to previous reports (7), our data suggest that individuals with low, or no, levels of neutralizing antibodies and total IgG antibodies have an increased risk of shedding live virus. Prior knowledge of severe acute respiratory syndrome coronavirus 1 (SARS-CoV-1) present similar data, where SARS-CoV-1 could be isolated during the first 2 weeks, but not after day 22, of illness (10). However, to better understand the correlation between virus shedding and serological response in COVID-19 patients, larger studies need to be performed.

The World Health Organization (WHO) recommends at least 10 days after symptom onset plus an additional 3 days without fever and respiratory symptoms, before releasing COVID-19 patients from isolation (11). Our data indicate that infectious SARS-CoV-2 could not be detected in the airways after day 14 after symptom onset despite detectable viral RNA by PCR unless immunocompromised patients. The latter finding is in concordance with a previous study also detecting viable viruses long after symptom onset in immunocompromised patients (12). Serological assay for SARS-CoV-2 may provide additional guidance in judging if a hospitalized patient may be shedding infectious SARS-CoV-2 virus, especially in the setting of immunocompromised COVID-19 patients.

## Material and methods

### Sample collection

Thirty-six patients with verified COVID-19 infection admitted to the Department of Infectious Diseases or the Intensive Care Unit at Karolinska University Hospital, Stockholm, Sweden, with a symptom duration >8 days and with a previous SARS-CoV-2 RNA positive nasopharynx or pharynx sample, were included in the study. One patient sampled 5 days after onset of symptoms was included as a positive control for virus shedding. Samples for culture and SARS-CoV-2 real time-PCR (RT-PCR) were collected from nasopharynx (all patients), and, when possible, sputum (30 patients). Serum were collected from all patients for SARS-CoV-2 RT-PCR analysis and antibody analysis.

### Ethical statement

The study was approved by the Swedish Ethical Review Authority, all relevant regulations for work with human participants were complied with, and patient samples were obtained according to the Declaration of Helsinki. Patients included in the study provided written informed consent.

### RT-PCR

SARS-CoV-2 RT-PCR were performed on respiratory samples and in serum. RT-PCR targeting the envelope(E)- and RNA dependent RNA polymerase (rdrp)-genes were used to detect the presence of SARS-CoV-2 RNA (13).

### Isolation of live SARS-CoV-2 from patient material

Virus isolation was carried out at a biosafety level 3 (BSL3) laboratory. For the virus cultures, 100 µL sample from nasopharyngeal swabs (NPS) and 100 µL tracheal secretion or sputum collected from a total of 37 patients, the positive control patient included, were inoculated in duplicate on Vero-E6 cells. Inoculation was carried out for 1.5 h at 37 °C and 5 % CO_2_ and medium (Dulbecco’s Modified Eagle’s Medium, DMEM) supplemented with 5 % fetal bovine serum (FBS), 10X Antibiotic-Antimycotic, 0.6 ug/mL penicillin, 60 ug/mL streptomycin, 2 mM L-glutamine, 20 mM HEPES) were then added. Cells were continuously monitored for cytopathic effect (CPE). After 10 days the cultures were harvested and supernatants were analyzed with RT-PCR specific for the SARS-CoV-2 E-gene and (rdrp)-gene (13).

### Sequencing of SARS-CoV-2

All primary clinical samples and samples from positive virus cultures were sequenced in order to exclude contamination. Sequencing libraries were constructed using the Ion AmpliSeq SARS-CoV-2 Research Panel (Thermo Fisher Scientific, MA, USA). cDNA was synthesized from SARS-CoV-2 RNA using SuperScript IV VILO Master Mix (Thermo Fisher Scientific, MA, USA) and incubated at 25 ℃ for 10 min, 50 ℃ for 10 min and 85 ℃ for 5 min. Automated library preparation and amplification was performed on an Ion Chef instrument with AmpliSeq Chef reagents and specific SARS-CoV-2 primers (Thermo Fisher Scientific, MA, USA). Libraries were quantified with TaqMan-based qPCR, using custom primers and probe targeting the flanking A- and P-adapters of the constructed libraries (Integrated DNA Technologies, IA, USA). The qPCR protocol was initiated by a holding stage at 95 ℃ for 20 s, followed by 40 cycles of 95 ℃ for 1 s and 60 ℃ for 20 s. Libraries were normalized to a final concentration of 35 pM and prepared for sequencing with the Ion 540 Kit-Chef (Thermo Fisher Scientific, MA, USA) for 200 base-pair reads. Sequencing was performed on an Ion GeneStudio S5 Sequencing System (Thermo Fisher Scientific, MA, USA). Raw reads were trimmed and filtered using default parameters, then analyzed with the Ion Torrent Suite software plugin coverageAnalysis, where an average read coverage of 20x for every amplicon of a sample was required to pass quality control. Variant calling against the SARS-CoV-2 reference sequence (NC_045512.2) was performed by the Ion Torrent Suite software plugin variantCaller.

### Immunofluorescence assay (IFA)

IFA was performed as previously described (13). Briefly, SARS-CoV-2-infected Vero cells mixed with uninfected Vero cells were used as target cells. Serum samples were heat-inactivated at 56°C for 30 minutes prior to analysis and two-fold serially diluted starting at 1:20. The titer of IgG in each serum sample was determined by the inverted dilution factor value for the highest dilution with positive staining.

### Micro-neutralization assay

IFA was performed as previously described (9). Heat inactivated (56°C for 30 minutes) serum were diluted in a two-fold dilution serie starting from 1:10. Each sample was prepared in duplicates. Each dilution was mixed with an equal volume of 4000 TCID_50_/ml SARS-CoV-2 (60 µl serum plus 60 µl virus). After incubation, 100 µL of the mixtures were added to Vero E6 cells seeded on 96-well plates and incubated at 37 °C 5% CO_2_. Four days later the cells were inspected for signs of cytopathic effect (CPE) by optical microscopy. Each well was scored as either ‘neutralizing’ if less than 50% of the cell layer showed signs of CPE, or ‘non-neutralizing’ if ≥50% CPE was observed.

## Data Availability

The data that support the findings of this study are available on request from the corresponding author (HG). The data are not publicly available due to restrictions e.g. their containing information that could compromise the privacy of research participants.

## Acknowledgements

The authors would like to thank the patients that contributed to the study. Further, authors would like to thank Karin Tegmark-Wisell and Shaman Muradrasoli at the Public Health Agency of Sweden for contributing to the design of the study. Parts of this study was supported by Marianne and Marcus Wallenberg Foundation (SGR), grants provided by region Stockholm (ALF project) (SGR), grants from the Swedish Research Council (ASS, JK) and the Swedish Heart and Lung Foundation (ASS).

## Authors contributions

H.G., S.G.R. and A.B. led the study.

M.O., M.L.K., S.B., J.K., and A.B. designed experiments and optimized assays.

H.G., S.F.J., R.F.J. and M.B. included patients and summarized clinical information.

M.O., K.T.M, W.C, M.L.K., S.B., and J.K. performed experiments.

H.G., S.G.R., M.O., S.F.J., R.V., W.C., K.T.M., M.L.K., S.B., N.J., A.F., A.S.S., J.K. and A.B. contributed to design and conceptualization of the study, discussed data analysis and interpreted the results.

H.G. and S.G.R wrote the paper, with input provided by all co-authors

## Conflict of interest

Authors declare no conflicts of interest.

## Notes

### Competing Interest Statement

The authors have declared no competing interest.

### Funding Statement

Parts of this study was supported by Marianne and Marcus Wallenberg Foundation (Sara Gredmark-Russ), grants provided by region Stockholm (ALF project) (Sara Gredmark-Russ), grants from the Swedish Research Council (Anna Smed-Sorensen, Jonas Klinstrom) and the Swedish Heart and Lung Foundation (Anna Smed-Sorensen).

### Author Declarations

Ethical approval, Sweden. Dnr: 2015/1949-31/4, Dnr: 2017/253-32, Dnr: 2018/382-32, Dnr: 2020-0055, Dnr: 2020-01757.

